# Optimization of TaqMan-based quantitative PCR diagnosis for *Entamoeba histolytica* using Droplet Digital PCR

**DOI:** 10.1101/2025.02.24.25322762

**Authors:** Akira Kawashima, Yasuaki Yanagawa, Takayuki Chikata, Rieko Shimogawara, Daisuke Mizushima, Kiyoto Tsuchiya, Kenji Yagita, Hiroyuki Gatanaga, Koji Watanabe

**Author notes:** Corresponding author Koji Watanabe, Department of Parasitology, Division of Host Defense Mechanism, Tokai University School of Medicine, 143 Shimokasuya, Isehara, Kanagawa, 259-1193, Japan.

## Abstract

**Background:** TaqMan-probed quantitative PCR (qPCR) is highly valued for diagnosing *Entamoeba histolytica* infections (amebiasis). However, unclear cycle threshold (Ct) values often yield low-titer positive results, complicating interpretation. This study aimed to optimize qPCR primer-probe sets with logically determined cut-off Ct value using droplet digital PCR (ddPCR).

**Methodology/Principal Findings:** Amplification efficacy was evaluated using ddPCR by measuring absolute positive droplet counts (APD) and mean fluorescence intensity at different PCR cycles and annealing temperatures (AT). A primer-probe specific cut-off Ct value was determined from a standard curve by correlating Ct values with APD. Twenty primer-probe sets targeting small subunit rRNA gene regions (X64142) were designed from previous papers. Amplification efficacy remained consistent at high PCR cycles (50 cycles), but differed at lower PCR cycles (30 cycles), identifying five sets with higher amplification efficiency than other candidates. Of these, only two sets maintained efficiency at higher AT (62°C). Ct value was inversely proportional to the square of APD, defining the specific cut-off Ct value as 36 cycles. Selected primer-probe set with a cut-off effectively differentiated *E. histolytica* infection in clinical specimens. However, discordant results between Ct value and APD were seen in some cases with high Ct value. Shotgun metagenomic sequencing suggested microbial-independent false-positive reactions contributed to these discrepancies, although specific reactants were unidentified.

**Conclusions/Significance:** The combination use of ddPCR with qPCR revealed that false positive reactions of qPCR and/or ddPCR commonly happen in stool specimens. Also, this study emphasizes the value of ddPCR for establishing accurate cut-off values with efficient primer-probes.

**Author Summary:** Accurate diagnosis of *Entamoeba histolytica* infection is critical for both clinical management and epidemiological monitoring. However, current diagnostic methods on TaqMan-qPCR for detecting this pathogen is hampered by inconsistent methodologies and ambiguous results, particularly in low-titer stool samples with high cycle threshold (Ct) values. To address this issue, this study introduces a novel approach using droplet digital PCR (ddPCR) to evaluate qPCR amplification efficacy and establish a robust theoretical cut-off Ct value for improved diagnostic accuracy. By evaluating different primer-probe sets, we identified optimal candidates and determined a Ct cut-off of 36 cycles, allowing for more reliable differentiation between true infections and false positives. Our findings enhance the diagnostic specificity and reliability, providing a practical and scalable framework for improving pathogen detection protocols, particularly in resource-limited settings. The methodology represents a significant advancement in molecular diagnostics, addressing a critical gap in accurate detection of *E. histolytica* in clinical stool samples and potentially other clinical specimens in clinical and research settings.

## Introduction

Amebiasis, transmitted orally, leads to diseases such as colitis and liver abscess ^1^. In developing countries, transmission typically occurs via the ingestion of food and water contaminated with feces, which is the second leading cause of parasite-related death worldwide ^2^. More recently, it has been reported that amebiasis is spreading throughout Asian and European developed countries as a sexually transmitted infection ^1,3–8^.

Among the currently available laboratory tests for *Entamoeba histolytica,* TaqMan-probed quantitative PCR (qPCR) is commonly used for clinical diagnosis and epidemiological studies because of its high sensitivity ^9^. However, the specificity of qPCR is still in doubt, especially when it is used for stool samples. We often face unexpected positive results with high Ct values in clinical settings, such as unexpected multiple pathogens detected by the intestinal panel of multiplex stool PCR system ^10^, and unexpectedly high frequency of asymptomatic carriers in epidemiological research ^11–13^. Previously, a high Ct was considered a feature of asymptomatic carriers rather than symptomatic diseases; however, recent studies identified the clinical severity of intestinal infection is not necessarily correlated with the Ct value ^14,15^. Taken together, the efficacy of qPCR should be assessed precisely, and cut-off Ct values should be set by logical strategy for better clinical practice.

Droplet digital PCR (ddPCR) is a third-generation PCR method that provides absolute quantification, enabling the partitioning of a sample into over 10,000 droplets, with each droplet serving as an independent reaction ^14,15^. The advantage of ddPCR is that fluorescence intensity is measured for each droplet, whereas qPCR measures the sum of the total template DNA in each sample ^16,17^. The fluorescence of each droplet represents an amplificon from one template DNA. The utility of ddPCR has been proven for clinical samples ^16^, although its use is still limited in the laboratory because of its cost and slightly complicated operation. In a study comparing ddPCR and qPCR, ddPCR accurately quantified samples, even at low concentrations, and it was less affected by contamination than qPCR ^18^. However, its utility has not been fully assessed in previous studies^17,20^.

The aim of this study was to optimize the diagnostic value of qPCR. For this, we newly applied ddPCR for evaluating the amplification efficacy of primer-probe sets, and determining cut-off Ct values by the logical approach.

## Methods

### Study participants and ethics

This study was conducted following ethical approval from the Institutional Review Board of the National Center for Global Health and Medicine (NCGM-S-004658). Written informed consent was obtained from all participants. Clinical samples were collected from patients with suspected *Entamoeba histolytica* infections.

### DNA Extraction

DNA was extracted from *E. histolytica* HM1:IMSS reference strains and clinical specimens using a QIAamp Fast DNA Stool Mini Kit (Qiagen, Hilden, Germany) according to the manufacturer’s protocol. Extracted DNA was stored at −20°C until analysis.

### ddPCR Protocol

Each reaction consisted of 10 μL ddPCR Master Mix for Probes (Bio-Rad, Munich, Germany), 18 pmol of each primer, 5 pmol of probes, and 1 μL DNA template adjusted to a final reaction volume of 20 μL. Droplets were generated with a QX200 Droplet Generator, transferred to a 96-well PCR plate, and amplified on a C1000 Touch™ Thermal Cycler under conditions of initial denaturation at 95°C for 10 minutes, followed by 20–50 cycles at 94°C for 30 seconds, 59–62°C for 1 minute, and a final extension at 98°C for 10 minutes. Amplified droplets were analyzed using a QX200 Droplet Reader to quantify absolute positive droplet counts (APD) and mean fluorescence intensity (MFI) using a Bio-Rad QX200 system.

### qPCR Protocol

Each 20 μL reaction contained 10 μL TaqMan Fast Advanced Master Mix 2× buffer (Thermo Fisher Scientific, Waltham, MA, USA), 5 pmol of each primer and probe, 2 μL template DNA and an internal positive control (Nippon Gene, Toyama, Japan) to assess potential PCR inhibition. Thermal cycling included an initial denaturation at 95°C for 30 seconds, followed by 40 cycles at 95°C for 5 seconds, and 59–62°C for 30 seconds. Ct values were analyzed using QuantStudio™ Design & Analysis software.

### Shotgun Metagenome Sequencing Analysis for Discordant PCR Cases

Shotgun metagenome sequencing was performed on discordant PCR results using the DNBSEQ platform. DNA libraries were prepared using a KAPA HyperPlus Kit according to the manufacturer’s protocol, with additional purification for high-quality DNA. Libraries were quantified with BioAnalyzer 2100 (Agilent Technologies) and sequenced as paired-end reads (2×150 bp).

Sequencing data were processed using a bioinformatics pipeline. Raw reads were subjected to adapter trimming, removal of low-quality sequences (limit = 0.05), and exclusion of reads shorter than 105 bp. Filtered reads were mapped to the *E. histolytica* reference sequences, including the small subunit rRNA gene (X64142) and HM-1:IMSS genome (JCVI-ESG2-1.0) using CLC Genomics Workbench with a length and similarity fraction threshold ≥ 0.8. Reads containing primer-homologous regions (allowing up to two mismatches) were identified and assembled *de novo* into contigs. Contigs were analyzed using BLAST against the NCBI core nt database to determine taxonomic origins based on significant alignments (E-values <10^5^).

### Statistical analysis

Statistical analyses were performed using GraphPad Prism (v10.2.3). The Pearson correlation coefficient was calculated to assess the relationship between APD counts and Ct values, with statistical significance defined as p < 0.05. Discrepancies were compared by one-way ANOVA followed by compact letter display for amplification efficiencies.

## Results

### Designing primer-probe sets for clinical specimens

For the clinical diagnosis of *E. histolytica,* primer-probe sets targeting the small subunit rRNA gene (X64142) were designed because of its high sensitivity. We designed twenty primer-probe sets in combination with primers and probes, all of which were proven for clinical utility in previous papers (Table 1 and 2). ^9,21–24^ Also, these primer-probe sets met the following criteria: 1. The size of the amplicon is < 300 bp for primers, and 2. Double quencher with ZEN enhancement can be designed for the probe.

**Table 1.**
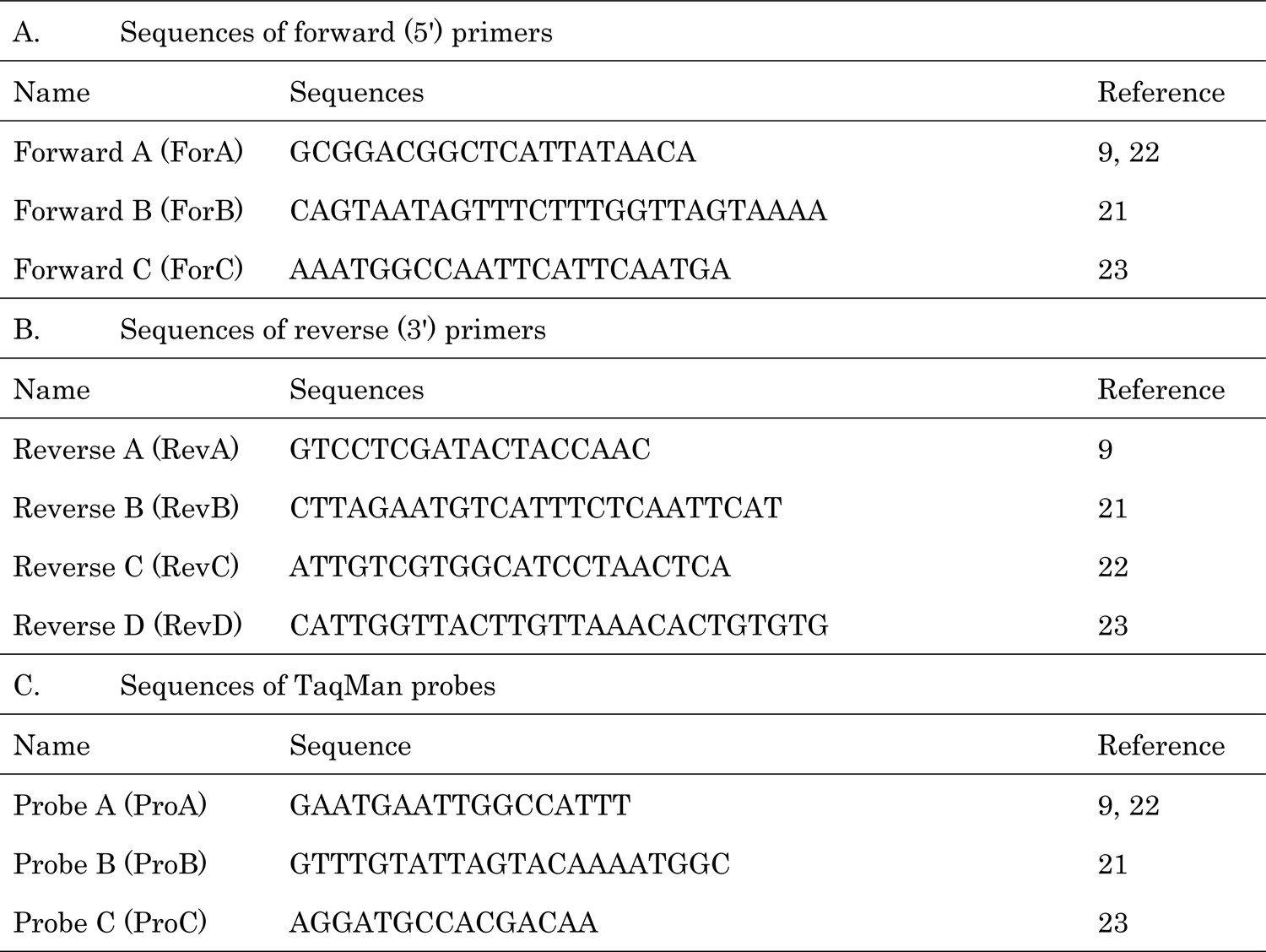
Sequences of primers used in this study.

### Comparison of amplification efficacy of each primer-probe set by ddPCR

#### Determination of highly sensitive primer-probe sets

First, we performed ddPCR to compare the amplification efficacy of each primer-probe set. We fixed the annealing temperature (59°C) and template concentration (1000 trophozoites per microliter) for the experiment. On the other hand, PCR cycles were decreased by 10 cycles from the standard number of PCR cycles (50 cycles). With 50 cycles, APD counts were similarly high in all primer-probe sets, moreover, a comparison of the MFI of positive droplets was difficult (Fig. 1A). Interestingly, as PCR cycles decreased, the differentiation of MFI became easier, although the MFI of all droplets decreased. When the cycle number was reduced to 30, APD counts were decreased in some primer-probe sets with relatively low MFI. Surprisingly, only five primer-probe sets (sets 3, 5, 12, 14, and 19) among 20 candidates maintained amplification efficacy, whose MFI levels were represented as “A” or “B” by the multiple comparison using GraphPad Prism© software (Fig. 1B).

**Figure 1.**
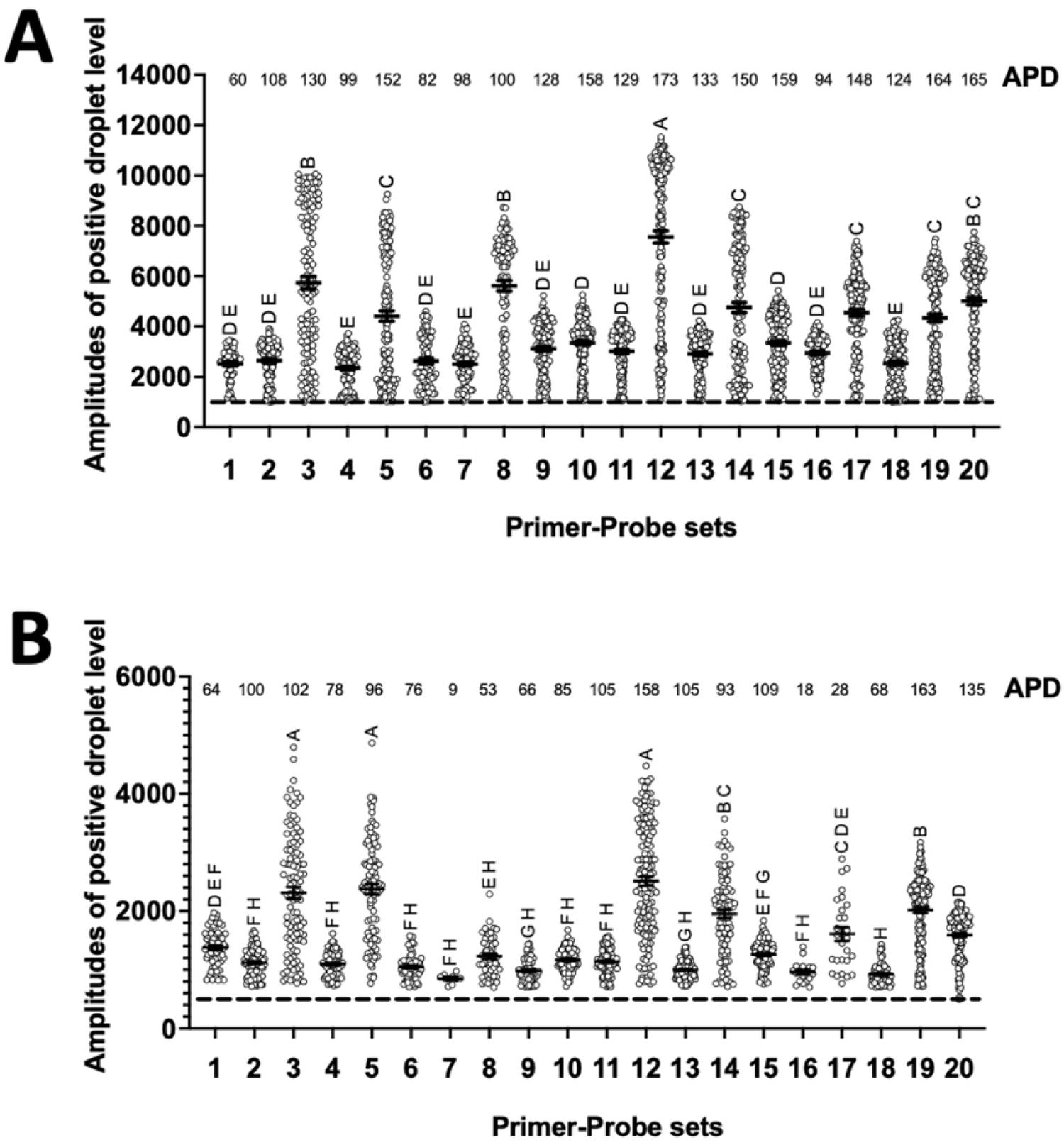
Evaluation of the amplification efficacy of primer-probe sets by droplet digital PCR (ddPCR). Absolute positive droplet counts (APD) and mean fluorescent intensity (MFI) were assessed by droplet digital PCR (ddPCR), which were compared among 20 prime-probe sets targeting the same *E. histolytica* gene (X64142). Assays were performed using different PCR cycles: 50 cycles (A), 40 cycles (B), 30 cycles (C), and 20 cycles (D). All conditions other than PCR cycles were the same for all experiments. MFIs were compared by one-way ANOVA and presented as compact letter display (CPD) using GraphPad Prism© software.

#### Determination of primer-probe sets with high specificity

Next, to identify primer-probe sets with low non-specific amplification in PCR, we compared the amplification efficacy at two different annealing temperatures, 59°C and 62°C (Fig. 2). Amplification efficacy was not decreased in sets 3 and 5, whereas that of the other sets was significantly decreased with a higher annealing temperature. Finally, to assess our ddPCR strategy for choosing effective primer-probe sets, we performed qPCR assay using these candidate primer-probe sets (Fig. 3A and B). As expected, sets 3 and 5 had lower Ct values compared with the other primer-probe sets, especially at higher annealing temperature. Sets 3 and 5 contain the same forward primer and probe, whereas the reverse primer is different (Table 2). Set 5 might amplify non-pathogenic *Entamoeba dispar,* possibly present in Japan, and the potentially pathogenic *E. moshkovskii*. Set 3 amplified *E. dispar*, and the different non-pathogenic *Entamoeba* species, *E. bangladeshii*. We selected set 5 for further evaluation in the present study with consideration of its future clinical use in Japan.

**Figure 2.**
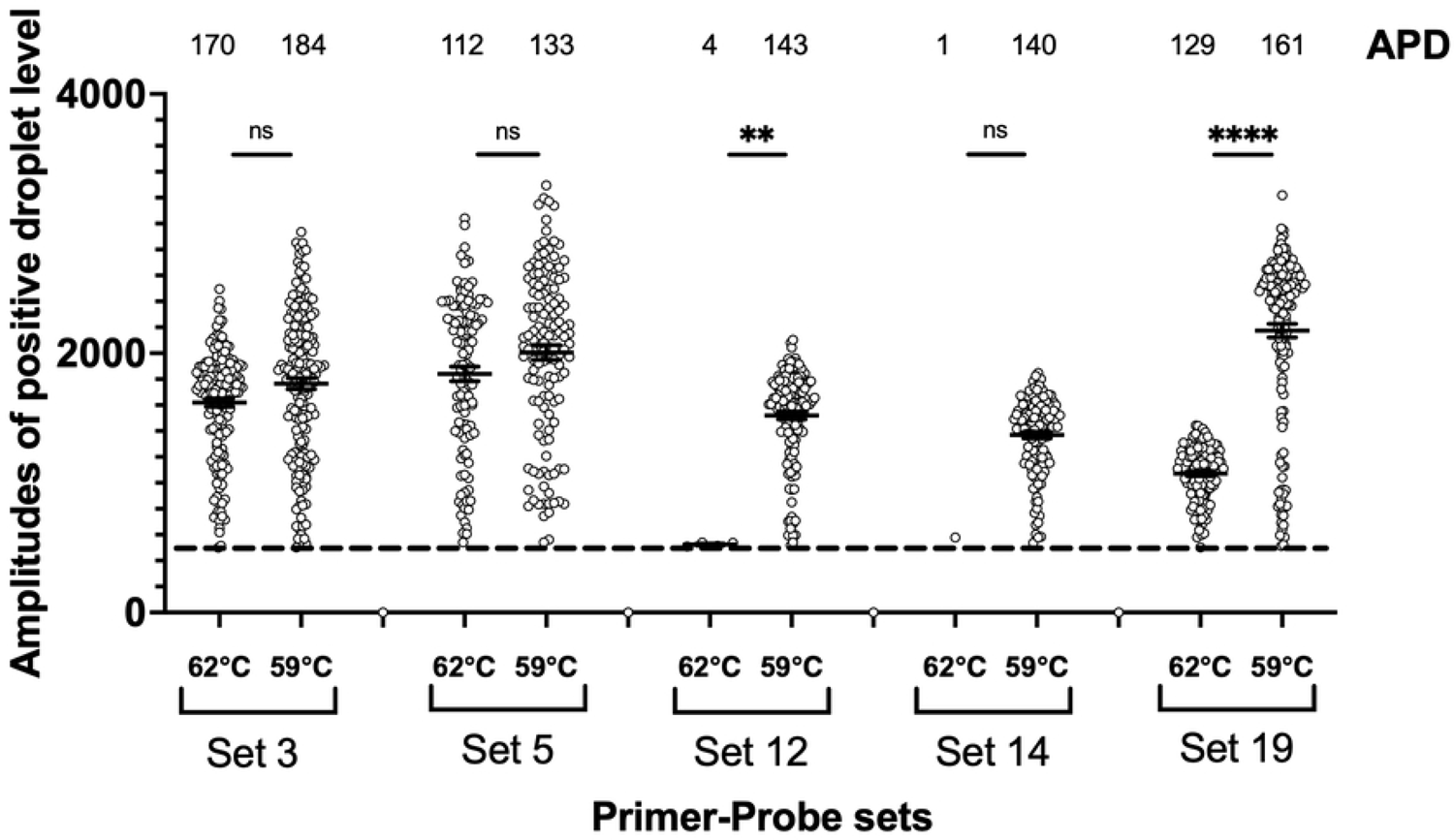
Evaluation of the amplification efficacy of five selected primer-probe sets at different annealing temperatures: 59°C versus 62°C.

**Figure 3.**
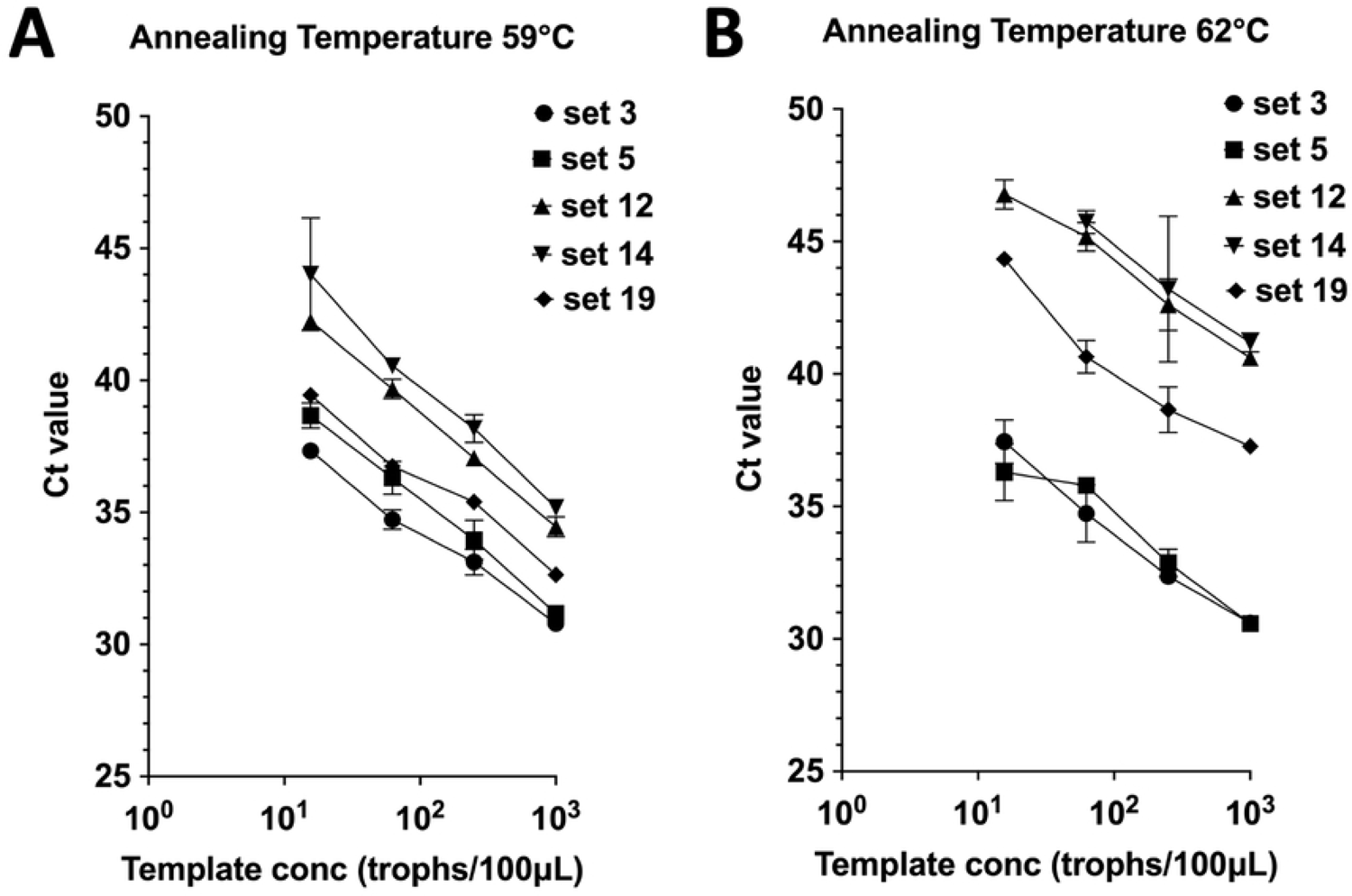
Evaluation of the diagnostic efficacy of TaqMan-probed quantitative PCR (qPCR). Ct values of five primer-probe sets were compared by the serial dilution of *E. histolytica* DNA. PCR was performed with 40 cycles at different annealing temperatures: 59°C (A), or 62°C (B).

**Table 2.** Primer-probe sets used in the present study.

| Primer-probe set | 5' Primer* | 3' Primer** | Amplicon length | Probe*** |
| --- | --- | --- | --- | --- |
| Set 1 | ForA | RevB | 151 bp | ProB |
| Set 2 | ForA | RevC | 173 bp | ProB |
| Set 3 | ForA | RevC | 173 bp | ProA |
| Set 4 | ForA | RevD | 207 bp | ProB |
| Set 5 | ForA | RevD | 207 bp | ProA |
| Set 6 | ForA | RevD | 207 bp | ProC |
| Set 7 | ForA | RevA | 247 bp | ProB |
| Set 8 | ForA | RevA | 247 bp | ProA |
| Set 9 | ForA | RevA | 247 bp | ProC |
| Set 10 | ForB | RevB | 135 bp | ProB |
| Set 11 | ForB | RevC | 157 bp | ProB |
| Set 12 | ForB | RevC | 157 bp | ProA |
| Set 13 | ForB | RevD | 191 bp | ProB |
| Set 14 | ForB | RevD | 191 bp | ProA |
| Set 15 | ForB | RevD | 191 bp | ProC |
| Set 16 | ForB | RevA | 231 bp | ProB |
| Set 17 | ForB | RevA | 231 bp | ProA |
| Set 18 | ForB | RevA | 231 bp | ProC |
| Set 19 | ForC | RevD | 99 bp | ProC |
| Set 20 | ForC | RevA | 139 bp | ProC |

\*The 5' primer was selected from Table 1A. \*\* The 3' primer was selected from Table 1B.
\*\*\*The probe was selected from Table 1C.

#### Determination of Cut-off Ct Values for TaqMan qPCR

Next, to determine cut-off Ct values specific to the primer-probe set, we applied ddPCR in combination with qPCR, which draws a standard curve between Ct value and APD. We performed ddPCR and qPCR using a series of different concentrations of *E. histolytica* HM-1 as templates and drew a standard curve between Ct value and APD (Fig. 4A). As expected, Ct values were inversely proportion to the square of APD counts (R^2^ = 0.9965, p value < 0.0001) with 95% confidence intervals (CI). Finally, we determined the primer-probe specific cut-off Ct value as 36 cycles, which is the detection limit of one positive droplet count in ddPCR.

**Figure 4.**
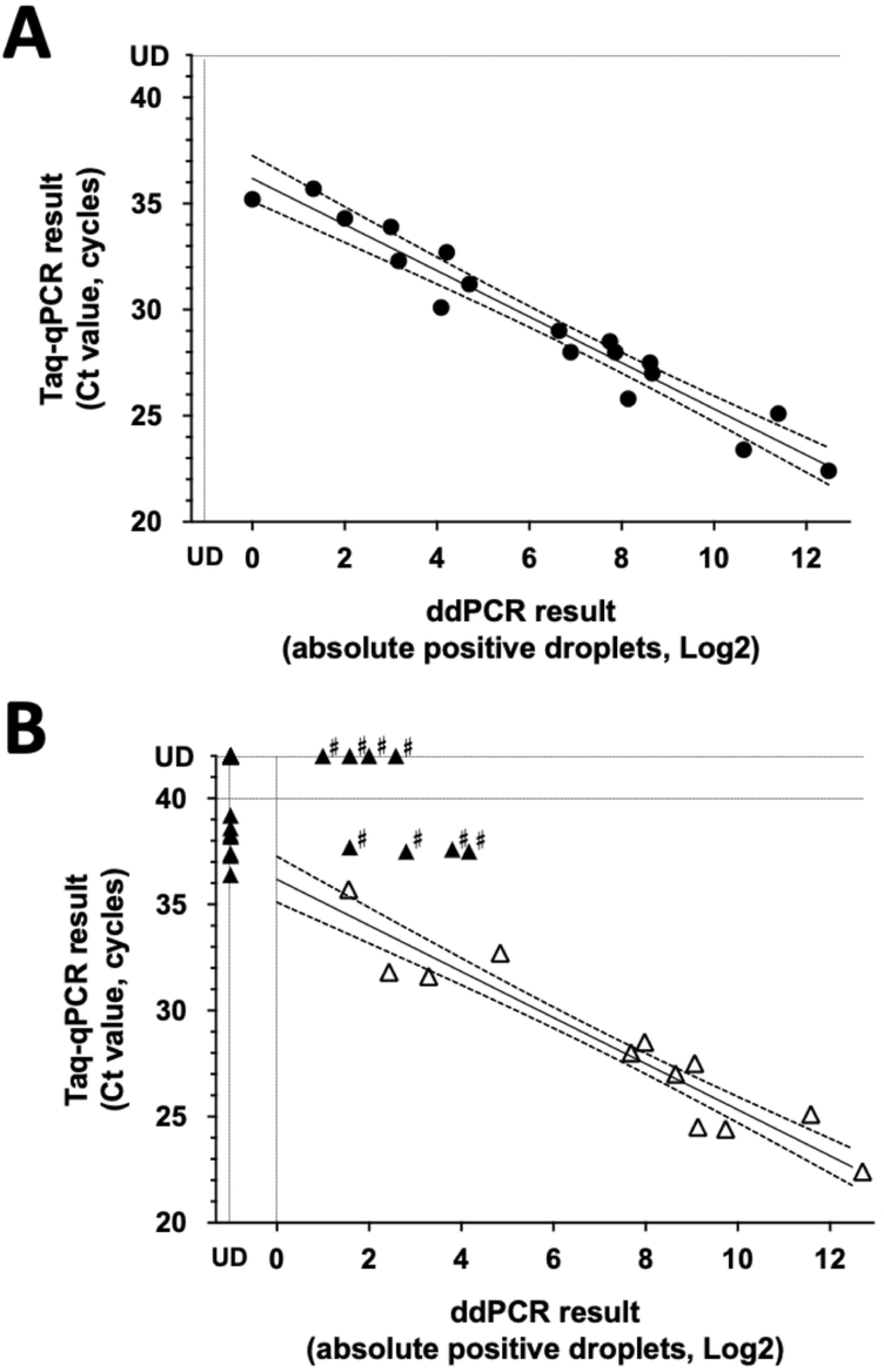
Primer-probe spefific Ct cut-off values for the laboratory strain of *E. histolytica* (A) and evaluation using clinical samples (B). A standard curve with 95% confidence intervals was drawn between Ct value by qPCR and square APD by ddPCR using a serial dilution of *E. histolytica* laboratory strain (R² = 0.9965, p < 0.0001). The primer-probe set 5 specific cut-off Ct value was determined as 36 cycles, which is the intersection of the standard curve on 1 APD by ddPCR (A). The APD and Ct value of clinical samples were directly plotted on the standard curve created in Fig. 4A. Open triangles represent the results of the samples with a lower Ct than the cut-off Ct value (Ct ≤ 36 by qPCR). Filled triangles represent the results of the samples with a higher Ct than the cut-off Ct value (Ct > 36, or undetermined by qPCR). The samples plotted far outside the 95% CI are indicated by # (discordant PCR results), some of which were transferred to the shotgun metagenome analysis to check the existence of *Entamoeba* derived DNA (B).

#### Performance of qPCR cut-off Ct value for clinical specimens

To assess the diagnostic value of primer-probe set 5 and the accuracy of the cut-off Ct value, we performed ddPCR and qPCR for 66 clinical samples with various Ct values (Supplementary data 1). We performed qPCR in duplicate and ddPCR four times for each sample to obtain reliable PCR results. Positivity was determined when positive results were obtained in all assays, and data was presented as the average titer.

For twelve samples with a lower Ct value than the cut-off Ct value (≤36), ddPCR was also positive. Moreover, APD was plotted around the 95%CI of the standard curve (represented as open triangles in Fig. 4B). Next, we examined ddPCR using 11 template DNA samples with positive reactions and higher Ct than the cut-off by qPCR assay (36<Ct<40 cycles). Positive droplets were not observed by ddPCR in seven cases, indicating these samples represented false positives by qPCR. Surprisingly, in the other four cases, ddPCR identified an unexpectedly high number of APD. Also, we applied ddPCR for 43 qPCR negative cases. Interestingly, ddPCR identified additionally identified an unexpectedly high number of APD in four qPCR negative cases (named as discordant PCR cases indicated by # in Fig. 4B).

Thus, ddPCR newly identified an unexpectedly high number of APD in some cases (discordant PCR cases). This indicated that fluorescence was generated in the PCR assay; however, fluorescence was not efficiently produced compared with that from the amplicon in the *E. histolytica* genome in these cases. To confirm the presence of the *Entamoeba* genome or other amplifiable organisms, we performed shotgun metagenome sequencing analysis for three discordant PCR cases (Table 3). However, analyses could not be performed for all cases, because the sample volume was not sufficient. We confirmed that the reads derived from the positive control (DDQ-55) were matched to the *E. histolytica* X16142 gene. We identified many reads from DDQ-55 were scattered to 36 different contigs of the *E. histolytica* genome of the *E. histolytica* reference strain (Table 3 and Supplementary data 2). However, reads derived from discordant PCR cases were never mapped to X16142 although some reads mapped to the *E. histolytica* whole genome. However, matched reads of these samples were mapped to the single contig. The sequence of the contig was the same as the other intestinal bacteria (99.8% similarity to *Clostridium perfringens* for NW_001915840, or 100% similarity to *Escherichia coli* for NW_001916323), indicating these reads were derived from these enteric bacteria, not from *Entamoeba spp*. Last, we examined sequences that might potentially match the forward or reverse primers. Many sequences from various intestinal organisms might match the primer sequences; however, the number of sequences was similar between the negative control and samples with discordant PCR cases. We could not identify the specific organism potentially amplified by the forward and reverse primer sets (Table 3 and Supplementary data 3).

**Table 3.** Read sequence analysis of the template solutions.

|  | DDQ-55<br>(Proven *E. histolytica*<br>positive control) | DDQ-47<br>(Proven *E. histolytica*<br>negative control) | DDQ-13 | DDQ-10 | DDQ-16 |
| --- | --- | --- | --- | --- | --- |
| Read information constructed from DNA library |  |  |  |  |  |
| Raw reads by DNB-SEQ | 26,352,760 | 38,636,884 | 38,606,267 | 39,019,871 | 24,169,710 |
| Filtered after excluding reads with low quality | 23,149,356 | 35,533,378 | 35,771,509 | 35,754,780 | 22,507,694 |
| Mapping results to *E. histolytica* small subunit<br>ribosomal RNA gene (X64142)* |  |  |  |  |  |
| Number of mapped reads to X64142 | 4 | 0 | 0 | 0 | 0 |
| BLAST search of mapped read sequence | 100% matched to *E.*<br>*histolytica* X64142 | N/A | N/A | N/A | N/A |
| Mapping results to *E. histolytica* whole genome of<br>HM1:IMSS* |  |  |  |  |  |
| Number of mapped reads | 67 | 0 | 13 | 2 | 25 |
| Number of mapped contigs of *E. histolytica* | 36 | N/A | 1*** | 1*** | 1**** |
| Potentially amplifiable reads by primer-probe set<br>5 |  |  |  |  |  |
| Primer matching organisms** (forward/reverse<br>primer) | 39/41 | 101/194 | 44/31 | 54/15 | 41/8 |

\*Conditions with length fraction (0.98) and similarity fraction (0.98) were applied. \*\*Less than 2 nucleotide mismatches were permitted for matching conditions.
\*\*\*All reads were mapped to the same contig (NW\_001915840, total length 1771 bp), which had 99.8% similarity to *Clostridium perfringens*. \*\*\*\*All reads were mapped to the same contig (NW\_001916323, total length 1031 bp), which had 100% similarity to *Escherichia coli* and other intestinal commensal bacteria.
Abbreviations: qPCR, quantitative PCR; UD, undetermined; BLAST, DNB-SEQ

Taken together, combination use of ddPCR with qPCR revealed that the false positive reactions can commonly happen in stool specimens as discordant PCR results. Also, it was suggested that the qPCR assay with logically determined cut-off Ct can confer the accurate diagnosis of *E. histolytica* infection in the clinical settings. However, the source of the non-specific fluorescence reaction causing false positive reactions was not identified in the present analyses.

## Discussion

The main objective of this study was the optimization of qPCR for the clinical diagnosis of *E. histolytica* infection. Standard ddPCR protocol with 50 cycles showed that sufficient numbers of positive droplets were obtained from all primer-probe candidates, regardless of amplification efficiency. However, with lower PCR cycles, only a few primer-probe sets obtained the same number of APD as those obtained with 50 cycles, moreover, differences in MFI between efficient and inefficient primer-probe sets were clearer with reduced cycles. Finally, we determined the two most effective primer-probe sets from an initial pool of 20 candidates. Next, we identified a primer-probe specific cut-off Ct value by plotting a standard correlation curve between Ct value and square APD. The utility of selected primer-probe set (set 5) and its cut-off Ct value were subsequently validated with clinical samples, which showing that this approach is practical for the clinical diagnosis. Of note, our strategy can be widely applied to other pathogens in the clinical specimen. Moreover, prior information on the target DNA concentration in the template is not required when determining cut-off Ct values, suggesting cut-off Ct values could be determined even for uncultivable pathogens using clinical specimens of unknown concentrations.

An additional objective of this study was to assess the significance of qPCR positivity with higher Ct values above the cut-off value, as these are frequently encountered in clinical practice and are often challenging to interpret. We analyzed clinical specimens using ddPCR and qPCR. Generally, positive droplets were not identified by ddPCR in cases with a high Ct by qPCR, which indicated that qPCR produced false positive results. Surprisingly, some samples exhibited an unexpectedly high number of APD by ddPCR in samples with undetermined or high Ct values by qPCR (discordant PCR cases). We performed shotgun metagenome sequencing analyses, which confirmed that *Entamoeba-*related DNA was not detected in these samples. Thus, we demonstrated that the false positive reactions can be commonly observed in stool specimen, which could be identified by the concomitant use of ddPCR and qPCR for the same sample. However, we could not identify the cause of these reactions. This study had some limitations. First, the cause of false positive reactions generated by qPCR and/or ddPCR in stool specimens could not be verified.

Experiments using different primer-probe sets that amplify the *Entamoeba* species or rRNA lesions of other organisms could not be used because of a limited source of clinical samples. Instead, we performed shotgun metagenome analysis; however, we could not identify the exact cause of the discordant results. Second, the sample composition in this study was predominantly stool samples with limited verification in other sample types. Future studies should extend the assay to include samples from other sources, such as liver abscesses and tissue biopsies, to confirm whether the calibration curve and cut-off values established here are applicable to a broader clinical context.

In conclusion, the combination use of ddPCR with qPCR revealed that false positive reactions of qPCR and/or ddPCR commonly happen in stool specimens. Also, this study emphasizes the value of ddPCR for establishing accurate cut-off values as well as assessing primer-probe efficiency for improving *E. histolytica* diagnosis in the clinical settings. Further studies should address non-specific fluorescent reactions in stools, and expand our strategy to a wider range of microbes and the other sample types for enhancing the diagnostic accuracy for infectious diseases.

## Data Availability

All data supporting the findings of this study will be made publicly available in appropriate repositories upon acceptance of the manuscript for publication. Due to the temporary system maintenance of DNA Data Bank of Japan (DDBJ, Accession number to be provided upon upload), data submission is currently unavailable. We will upload our raw sequencing data as soon as the database recovers, expected by late March 2025. Additional data may be available upon reasonable request.

## Acknowledgments

We are grateful to Shosei Kubota at Fasmac Co., Ltd. and the staff at the AIDS Clinical Center at the National Center for Global Health and Medicine for their contribution to the present study. We thank J. Ludovic Croxford, PhD, from Edanz (https://jp.edanz.com/ac) for editing a draft of this manuscript. This work was supported by the Emerging/Re-emerging Infectious Diseases Project of Japan from the Japan Agency for Medical Research and Development (grant numbers JP24jk0210050h0001, JP24fk0108681h0702 and JP24fk0108680s0202), and a grant from the National Center for Global Health and Medicine (23A2017).

All authors significantly contributed to this study. Study design and conceptualization were conducted by A.K. and K.W., who were overall responsible for the study. A.K. and K.W. wrote the manuscript. A.K., R.S., and Kenji Yagita K.Y. optimized the PCR analysis and developed the ddPCR methodology. Takayuki T.C. and K.W. verified the NGS data. A.K. and Y.Y. recruited participants and managed clinical data collection. D.M., K.T., and H.G. supervised the study. Figures and tables were prepared by A.K. and K.W. Funding acquisition was performed by A.K. and K.W. All authors have approved the final version of the manuscript.

---

**Supplementary Data 1.** Sample information of clinical specimens.

**Supplementary Data 2.** Contig information of *E. histolytica* to which reads derived from clinical samples were mapped.

**Supplementary Data 3.** Read sequences of other intestinal organisms that were potentially amplified by primer set 5.

